# Plasma-based antigen persistence in the post-acute phase of SARS-CoV-2 infection

**DOI:** 10.1101/2023.10.24.23297114

**Authors:** Michael J. Peluso, Zoe N. Swank, Sarah A. Goldberg, Scott Lu, Thomas Dalhuisen, Ella Borberg, Yasmeen Senussi, Michael A. Luna, Celina Chang Song, Alexus Clark, Andhy Zamora, Megan Lew, Badri Viswanathan, Beatrice Huang, Khamal Anglin, Rebecca Hoh, Priscila Y. Hsue, Matthew S. Durstenfeld, Matthew A. Spinelli, David V. Glidden, Timothy J. Henrich, J. Daniel Kelly, Steven G. Deeks, David R. Walt, Jeffrey N. Martin

## Abstract

**BACKGROUND:** Persistent symptoms among some persons who develop COVID-19 has led to the hypothesis that SARS-CoV-2 may, in some form or location, persist for long periods following acute infection. Several studies have shown data in this regard but are limited by non-representative and small study populations, short duration since acute infection, and lack of a true-negative comparator group to assess assay specificity.

**METHODS:** We evaluated adults with RNA-confirmed COVID-19 at multiple time points following acute infection (pandemic-era participants) and adults with specimens collected prior to 2020 (pre-pandemic era). Using once-thawed plasma, we employed the Simoa® (Quanterix) single molecule array detection platform to measure SARS-CoV-2 spike, S1, and nucleocapsid antigens.

**RESULTS:** Compared to 250 pre-pandemic participants who had 2% assay positivity, detection of any SARS-CoV-2 antigen was significantly more frequent among 171 pandemic-era participants at three different time periods in the post-acute phase of infection. The absolute difference in SARS-CoV-2 plasma antigen prevalence was +11% (95% CI: +5.0% to +16%) at 3.0-6.0 months post-onset of COVID-19; +8.7% (95% CI: +3.1% to +14%) at 6.1 to 10.0 months; and +5.4% (95% CI: +0.42% to +10%) at 10.1-14.1 months. Hospitalization for acute COVID-19 and, among the non-hospitalized, worse self-reported health during acute COVID-19 were associated with greater post-acute phase antigen detection.

**CONCLUSIONS:** Compared to uninfected persons, there is an excess prevalence of SARS-CoV-2 antigenemia in SARS-CoV-2-infected individuals up to 14 months after acute COVID-19. These findings motivate an urgent research agenda regarding the short-term and long-term clinical manifestations of this viral persistence.

## BACKGROUND

Early in the COVID-19 pandemic, there were case reports of prolonged detection of SARS-CoV-2 RNA, well past the period of acute symptoms, in surface samples from the nasopharynx and gastrointestinal tract, two sites known to be involved in primary infection. Systematic investigation, however, revealed RNA detection after 28 days following symptom onset in immunocompetent hosts was uncommon and rarely documented to exceed 90 days.^1–7^ More recently, there has been detection of either SARS-CoV-2 RNA or protein in other sites, such as blood or gastrointestinal tract tissue, at durations of three months or more following acute infection.^8^ These findings suggest a phenomenon beyond that of the residual tail of primary viral infection; they suggest a persistent SARS-CoV-2 reservoir. Given the sheer number of persons who have been infected with SARS-CoV-2, a persistent reservoir of virus — even if clinically relevant in a small percentage of persons — could have substantial implications at the population level.

While intriguing, the initial data supporting persistent SARS-CoV-2, in some form or location, are short of definitive. Use of small and non-representative sample sizes, unclear documentation of complicating vaccination and reinfection histories, and, most importantly, lack of a sizeable true-negative comparator group to assess assay specificity all threaten conclusions about the existence, frequency, and duration of viral persistence.^9–16^ To address these limitations, we evaluated the presence of SARS-CoV-2 antigens in plasma in a large, well-characterized group of persons at several time points in the first 14 months of the post-acute phase of SARS-CoV-2 infection, most of whom were studied prior to widespread vaccination or reinfection. To understand the specificity of our findings, we compared them to a large number of persons studied prior to 2020, who, by definition, were SARS-CoV-2-uninfected. Finally, we evaluated several host and acute COVID-19-related factors for their influence on persistent SARS-CoV-2 antigenemia.

## METHODS

### Overall Design

In cross-sectional analyses, we compared participants in the post-acute phase of SARS-CoV-2 infection to persons studied prior to the COVID-19 pandemic for the presence of three SARS-CoV-2 antigens in plasma. Among persons in the post-acute phase of SARS-CoV-2 infection, we also evaluated several sociodemographic characteristics and clinical factors related to acute COVID-19 for their influence on SARS-CoV-2 antigen detection in the post-acute phase.

### Participants

We studied two groups of participants. The first (hereafter known as pandemic-era) were participants in the University of California, San-Francisco (UCSF)-based Long-term Impact of Infection with Novel Coronavirus (LIINC) study (NCT04362150). Selection of participants has been described previously.^17^ Briefly, using facility- and community-based advertising (through the internet and word-of-mouth), we enrolled (beginning in April 2020) consecutive adult volunteers who had earlier experienced their first episode of acute SARS-CoV-2 infection (confirmed by detection of SARS-CoV-2 RNA or antigen) and who were at least two weeks removed from their onset of symptoms. These volunteers were responding to advertising that described the study’s interest in a variety of long-term biochemical and clinical outcomes of COVID-19. Participants were examined at an initial study visit and every four months thereafter. For the present analysis, we sampled participants who had the greatest number of completed study visits (with stored plasma specimens) in the first 1.25 years following COVID-19 onset. The second group of participants (hereafter known as pre-pandemic-era) were from the UCSF-based Study of the Consequences of the Protease Inhibitor Era (SCOPE), a cohort study begun in 2001 originally focused on pathogenesis of HIV infection. It contains participants at variety of stages of HIV infection as well as ambulatory HIV-uninfected comparators, all of whom were volunteers from the community. For the present analysis, we randomly selected, among SCOPE participants with stored plasma specimens prior to December 2019, four HIV-uninfected participants to every one HIV-infected participant, attempting to match the age and race/ethnicity distribution of the pandemic-era group. All participants provided written informed consent.

### Measurements

Questionnaire-based. In both groups, interviewer-administered questionnaires collected data on sociodemographic and economic characteristics. In the pandemic-era group, we also inquired about details concerning the acute phase (first 3 weeks) of SARS-CoV-2 infection, including symptoms experienced, self-reported worst perception of overall health on a 0 to 100 scale, and whether hospitalization for COVID-19 occurred. The pandemic-era group also had all SARS-CoV-2 vaccinations recorded as well as any additional SARS-Co-V-2 re-infections since the initial infection.

Laboratory-based. Peripheral blood was collected in EDTA-coated tubes and plasma stored at −80° C using similar procedures in both the pre-pandemic era and pandemic-era groups. Using once-thawed plasma, we employed the Simoa® (Quanterix) single molecule array detection platform to measure SARS-CoV-2 antigens from spike, S1, and nucleocapsid (N) proteins; detailed methods have been described elsewhere.^13,18^ Briefly, plasma samples were centrifuged at 2000 x g for 10 minutes at 4° C and treated with 5 mM dithiothreitol (Pierce^TM^ No-Weigh^TM^ Format, Thermo Fisher Scientific) and protease inhibitors (Halt^TM^ Protease Inhibitor Cocktail, Thermo Fisher Scientific) for 15 minutes at 37° C. Each plasma sample was diluted 8-fold in a 96-well plate with Sample Diluent Buffer (Quanterix) and analyzed automatically with a three-step format on a HD-X Analyzer (Quanterix). In the first step, the plasma samples are incubated with antibody-coated magnetic beads. Assays for S1, spike, and N were performed separately, using antibodies against S1 (40150-D006, Sino Biological), S2 (MA5-35946, Invitrogen), and N (40143-R004, Sino Biological) conjugated to carboxylated magnetic beads (Quanterix). In the second step, the beads are resuspended in a solution of biotinylated detector antibodies. The same detector antibody against S1 is used for the S1 and spike assays (LT-1900, Leinco) and another antibody against N is used for the N assay (40143-R040, Sino Biological). In the third step, the beads are incubated in a solution of streptavidin conjugated β-galactosidase and lastly resuspended in a solution of resorufin β-D-galactopyranoside and loaded into a microwell array. The array is then sealed with oil and imaged. Average enzyme per bead (AEB) values are calculated by the HD-X Analyzer software thereafter and converted to concentration values based on a calibration curve fit with a four-parameter logistic regression. Separately, the limit of detection (LOD) is calculated as the background AEB plus three times the standard deviation and converted to a concentration. The LOD was determined to be 14.47 pg/mL for the spike assay, 11.16 pg/mL for S1, and 4.55 pg/mL for N.

### Statistical analysis

When comparing antigen prevalence in the pandemic-era group to the pre-pandemic era group, we defined three time periods for the pandemic-era group: 3.0-6.0 months, 6.1-10.0 months, and 10.1-14.1 months post-onset of COVID-19 symptoms. If there was more than one time point per person in a given time period, we chose the timepoint closest to the period’s midpoint. In each plasma specimen tested, antigen detection was defined in four ways: a) presence or absence on each of three individual antigen assays; and b) presence of at least one of the three antigens (vs absence on all three). Comparison between groups were expressed with prevalence ratios and differences. All analyses were performed using Stata version 17.0 (StataCorp, College Station, Texas).

## RESULTS

### Study participants

We studied 171 pandemic-era participants, who contributed 660 plasma specimens obtained between 0.9 and 14.1 months following initial SARS-CoV-2 symptom onset, and 250 pre-pandemic-era participants who each contributed one plasma specimen between 2003 and 2019 (Table 1). The groups were similar in age, but the pandemic-era group had more women, more Latino and White participants, and higher measures of socioeconomic status. Both groups originated from underlying research studies that were deliberatively enriched for people with HIV (PWH), and, as a result, the prevalence of HIV infection was similar in the two groups but much higher than the general population. All but four participants in pandemic-era group developed COVID-19 prior to SARS-CoV-2 vaccination and prior to the Omicron era.

**Table 1.** Characteristics of pre-pandemic-era and pandemic-era participants who were examined for the presence of SARS-CoV-2 antigens in plasma.

| Characteristic | Pandemic Era<br>(n = 171) | Pre-Pandemic Era<br>(n = 250) |
| --- | --- | --- |
| **Age, years\*** | 46 (37-57) | 48 (36-58) |
| **Female Birth Sex** | 86 (50%) | 56 (22%) |
| **Sexual Orientation<sup>†</sup>** |  |  |
| Asexual | 1 (0.58%) | 0 (0.0%) |
| Bisexual | 1 (0.58%) | 24 (9.6%) |
| Gay/lesbian | 34 (20%) | 98 (39%) |
| Straight/heterosexual | 110 (64%) | 113 (45%) |
| Questioning/unsure | 3 (1.8%) | 0 (0.0%) |
| Other | 1 (0.58%) | 0 (0.0%) |
| **Race/Ethnicity<sup>†</sup>** |  |  |
| Hispanic/Latino | 47 (28%) | 41 (16%) |
| White | 92 (54%) | 104 (42%) |
| Black/African American | 8 (4.7%) | 76 (30%) |
| Asian | 17 (9.9%) | 29 (12%) |
| Pacific Islander/Native Hawaiian | 3 (1.8%) | 0 (0.0%) |
| **Education<sup>†</sup>** |  |  |
| Any high school or less | 33 (19%) | 79 (32%) |
| Any college | 69 (40%) | 117 (47%) |
| Any graduate school | 69 (40%) | 52 (21%) |
| **Income<sup>†</sup>** |  |  |
| \$30,000 or less | 24 (14%) | 115 (46%) |
| \$30,001 to \$70,000 | 24 (14%) | 36 (14%) |
| More than \$70,000 | 99 (58%) | 40 (16%) |
| **Body Mass Index<sup>†</sup>** |  |  |
| Less than 18.5 kg/m<sup>2</sup> | 2 (1.2%) | 5 (2.0%) |
| 18.5 kg/m<sup>2</sup> to 24.9 kg/m<sup>2</sup> | 58 (34%) | 116 (46%) |
| 25.0 kg/m<sup>2</sup> to 29.9 kg/m<sup>2</sup> | 49 (29%) | 68 (27%) |
| More than 30.0 kg/m<sup>2</sup> | 61 (36%) | 50 (20%) |
| **HIV Seropositive** | 25 (15%) | 50 (20%) |
| **Hospitalized During Acute COVID-19 Infection<sup>†</sup>** | 33 (19%) | N/A |
| **Symptom Count During Acute COVID-19 Infection\*** | 9 (6-12) | N/A |
| **Self-reported Health at Worst Point in Acute COVID-19<sup>\*‡</sup>** | 45 (25-60) | N/A |
| **Self-reported Current Health<sup>†</sup>** |  |  |
| Excellent | N/A | 62 (25%) |
| Very Good | N/A | 82 (33%) |
| Good | N/A | 65 (26%) |
| Fair | N/A | 26 (10%) |
| Poor | N/A | 3 (1.2%) |
| **Time from COVID-19 Symptom Onset to Enrollment, days\*** | 56 (37-85) | N/A |

\*Median (interquartile range); <sup>†</sup>Missing and nonresponse. Sexual orientation: 35 missing, 1 prefer not to answer; Race/ethnicity: 4 missing; Education: 2 missing; Income: 3 missing, 80 prefer not to answer; BMI: 12 missing; Hospitalization: 1 missing; Self-reported health score at worst point in COVID illness: 82 missing; Self-reported current health: 12 missing. <sup>‡</sup>Response to "Using a scale from 0 to 100, we would like to know how good or bad your health was during the time you had COVID-19. A score of 100 means the best health you can imagine, and 0 means the worst health you can imagine." N/A denotes not asked or not applicable.

### SARS-CoV-2 antigen detection in pandemic era compared to pre-pandemic era

Positivity in at least one SARS-CoV-2 antigen assay was present in the plasma of 5 pre-pandemic participants (2%; 95% confidence interval (CI): 0.65% to 4.6%). Given the definitional absence of target in these specimens, these 5 instances are considered false positive, thus establishing any-antigen assay specificity to be 98% (95% CI: 95% to 99%). For the individual antigens, spike was detected in 3 (1.2%) participants, S1 in 3 (1.2%), and N in 2 (0.80%). The 5 participants were a 20-to-29-year-old White woman (S1 alone [34.67 pg/mL]), 50-to-59-year-old Asian man (spike alone [83.46 pg/mL]), 40-to-49-year-old White man (spike alone [609.96 pg/mL]), 30-to-49-year-old Asian man (S1 [285.31 pg/mL] and N [649.28 pg/mL]), and 40-to-49-year-old White woman (spike [646.02 pg/mL], S1 [115.89 pg/mL], and N [5716.32 pg/mL]). Of the 5, 2 (40%) were HIV-seropositive.

Of the 660 pandemic-era plasma specimens tested, 61 (9.2%) representing 42 unique participants (25% of the group) had one or more detectable SARS-CoV-2 antigens (Figure 1). The most commonly detected antigen was spike (n=33, 5.0%), followed by S1 (n=15, 2.3%) and N (n=15, 2.3%). In most instances (59/61, 97%) only a single antigen was detected; one specimen was positive for S1 and N and a second was positive for spike and N. Of those with detectable antigen, the median (IQR) concentration was 27.7 pg/mL (IQR 20.5 to 33.7) for spike, 31.2 pg/mL (IQR 20.5 to 193.0) for S1, and 23.6 pg/mL (IQR 6.46 to 62.0) for N.

**Figure 1.**
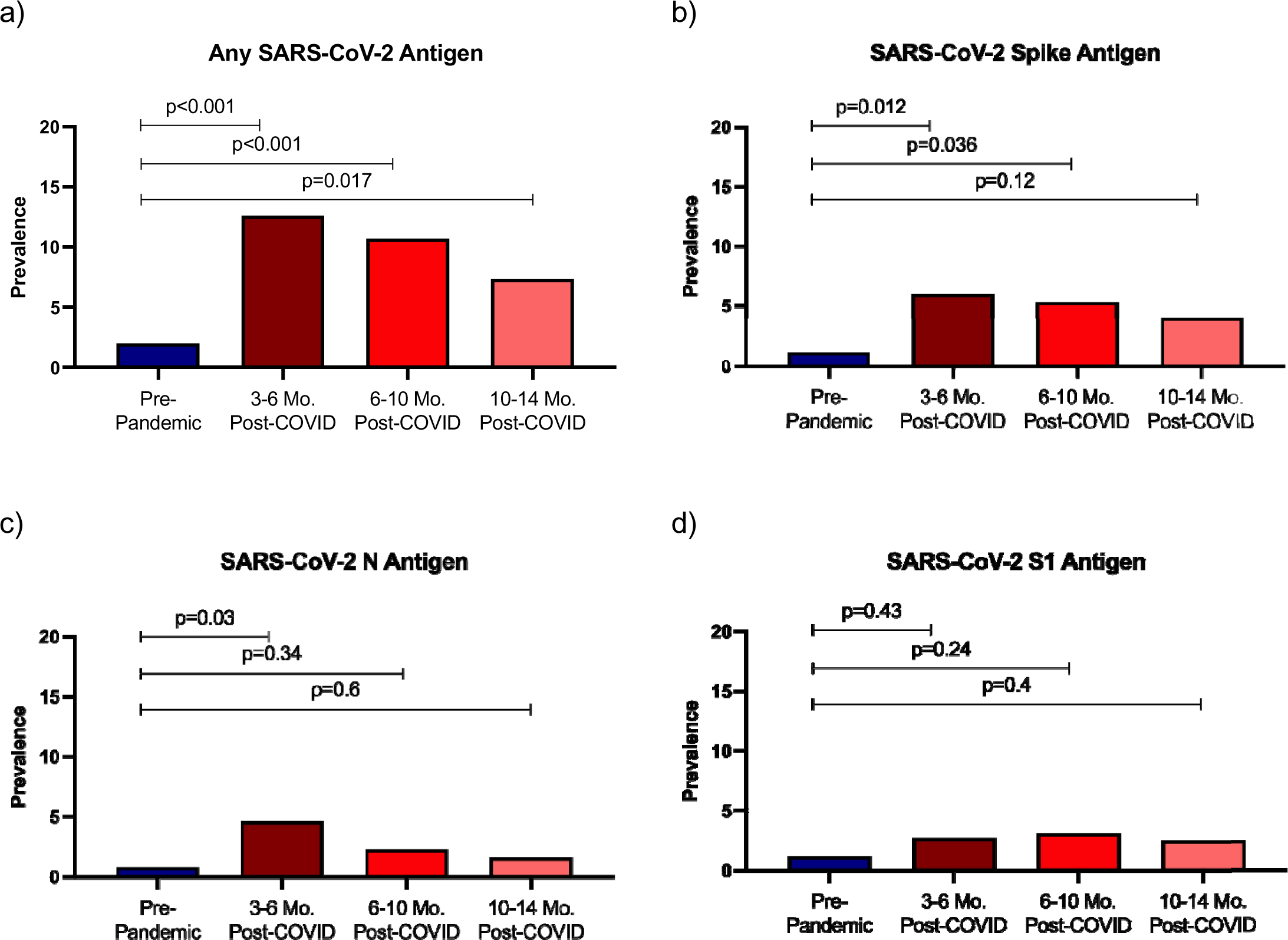
Prevalence of SARS-CoV-2 antigen positivity in plasma among participants in the post-acute phase of COVID-19 in comparison to pre-pandemic participants. P-values represent chi-square and Fisher’s 2-sided exact test as appropriate. (a) Prevalence of any SARS-CoV-2 antigen positivity (Spike, S1 or nucleocapsid); (b) Spike antigen prevalence; (c) Nucleocapsid antigen presence; and (d) S1 antigen presence.

Compared to the pre-pandemic group, detection of any SARS-CoV-2 antigen was significantly more frequent among pandemic era participants at all three time periods that we evaluated in the post-acute phase of infection. The absolute difference in antigen prevalence was +11% (95% CI: +5.0% to +16%) at 3.0-6.0 months post-onset of COVID-19; +8.7% (95% CI: +3.1% to +14%) at 6.1 to 10.0 months; and +5.4% (95% CI: +0.42% to +10%) at 10.1-14 months (Figure 2a; Supplemental Table 1). Regarding the individual antigens, significant differences between pandemic-era and pre-pandemic-era participants were observed for spike protein for up to 10 months and for N in the first 6 months after infection (Figure 2b-d).

**Figure 2.**
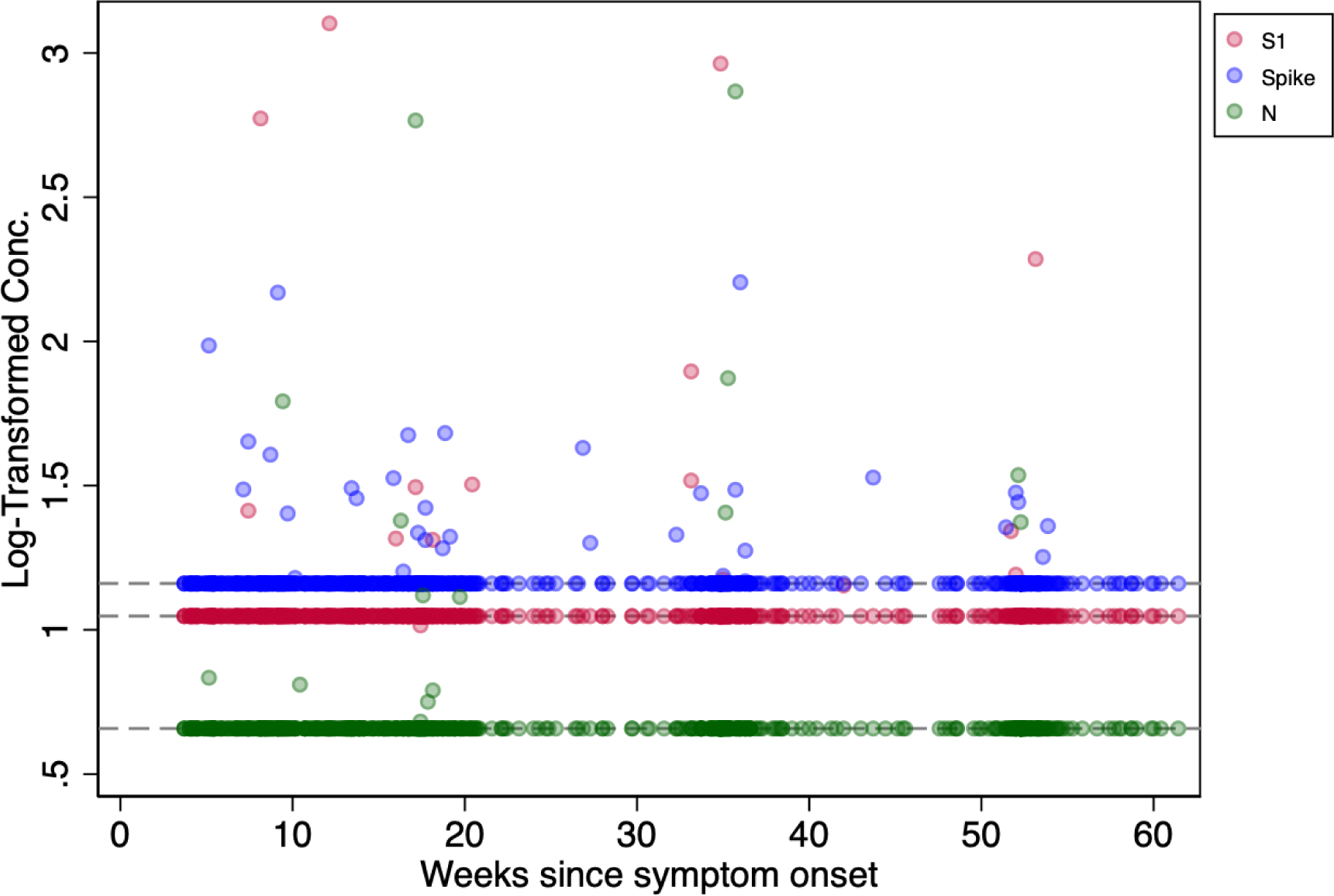
Quantitative measurement of SARS-CoV-2 spike, S1, and nucleocapsid antigens in plasma among all pandemic-era participants during the post-acute phase of SARS-CoV-2 infection. Dashed lines indicate limit of quantification for each antigen. Y-axis refers to log-transformed concentration of antigen in picograms per mL.

### Profiles of SARS-CoV-2 antigen positivity over time within individual pandemic-era participants

Of 159 pandemic-era participants who had multiple timepoints studied, 29 (18%) had antigen detected at a single post-acute timepoint, 10 (6.3%) had antigen detected at two post-acute timepoints, and one (0.63%) had antigen detected at three, four, and five post-acute timepoints, respectively (Figure 3). Most timepoints at which antigen was detected (51/61, 84%) occurred before the participant had ever received a SARS-CoV-2 vaccine. There were five instances in which antigen was detected within three weeks of a SARS-CoV-2 vaccine dose (three for S1, one for Spike, and one for N).

**Figure 3.**
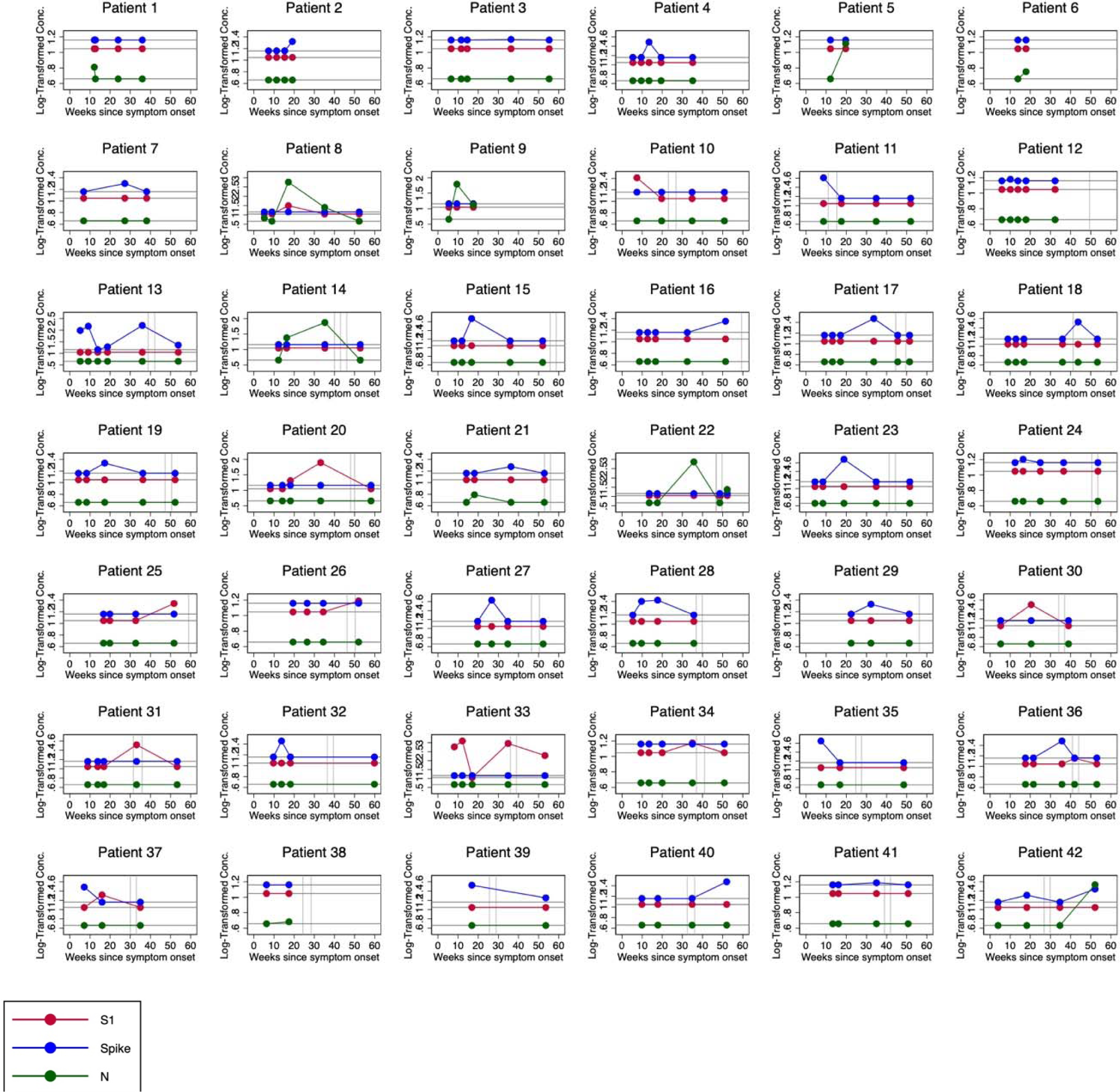
Individual participant-level profiles of quantitative measurement of SARS-CoV-2 spike, S1, and nucleocapsid antigens in plasma during the post-acute phase of SARS-CoV-2 infection, limited to participants with at least one positive antigen. Blue indicates spike, green nucleocapsid, and red S1. Horizontal dotted lines represent the assay limit of detection for each antigen. Vertical dotted lines indicate receipt of SARS-CoV-2 vaccine. Y-axis refers to log-transformed concentration of antigen in picograms per mL.

### Determinants of antigen positivity among pandemic-era participants

Among the pandemic-era participants, we found no strong evidence of an association between age, sex, race/ethnicity, HIV status, or body mass index (BMI) with SARS-CoV-2 antigen positivity at any point between 3 and 14 months in the post-acute period of infection (Table 2). In contrast, we found several markers of severity of the acute phase of SARS-CoV-2 infection to be related to subsequent SARS-CoV-2 antigen positivity in the post-acute phase. As compared to those not hospitalized, participants who required hospitalization for acute COVID-19 were nearly twice as likely to have antigen detected (prevalence ratio [PR] 1.97; 95% CI: 1.11 to 3.48), an absolute difference of +18% (95% CI: 0% to +37%). Among those not hospitalized for COVID-19, those who reported the worst overall health during the acute phase of COVID-19 (on a 0 to 100 scale) were over 2.5 times as likely to have antigen detected as compared to those with the most benign self-report (PR 2.82; 95% CI: 0.66 to 12.1), an absolute difference of +23% (95% CI: −5.0% to +51%).

**Table 2.** Association between sociodemographic and clinical characteristics and SARS-CoV-2 antigen positivity (detection of spike, S1, or nucleocapsid antigen at any timepoint 3 to 14.1 months post-COVID-19 onset) among pandemic-era participants.

| Characteristic | Prevalence | Prevalence Ratio (95% CI) | Prevalence Difference (95% CI) | P value |
| --- | --- | --- | --- | --- |
| **Age, years** |  |  |  |  |
| Less than 40 | 0.26 | Ref | Ref |  |
| 41-65 | 0.19 | 0.71 (0.38 to 1.31) | -0.08 (-0.22 to +0.07) | 0.28 |
| Greater than 65 | 0.33 | 1.26 (0.54 to 2.95) | +0.07 (-0.20 to +0.34) | 0.60 |
| **Sex at Birth** |  |  |  |  |
| Female | 0.17 | Ref | Ref |  |
| Male | 0.28 | 1.68 (0.93 to 3.04) | +0.12 (-0.01 to +0.24) | 0.081 |
| **Race/ethnicity\*** |  |  |  |  |
| White | 0.21 | Ref | Ref |  |
| Hispanic/Latino | 0.32 | 1.51 (0.83 to 2.72) | +0.11 (-0.05 to +0.27) | 0.18 |
| Black/African American | 0.25 | 1.12 (0.33 to 4.21) | +0.04 (-0.27 to +0.35) | 0.80 |
| Asian | 0.12 | 0.59 (0.15 to 2.31) | -0.09 (-0.75 to +0.10) | 0.43 |
| **HIV Infection Status** |  |  |  |  |
| Uninfected | 0.23 | Ref | Ref |  |
| Infected | 0.21 | 0.91 (0.39 to 2.11) | -0.02 (-0.20 to +0.16) | 0.83 |
| **Body Mass Index\*** |  |  |  |  |
| 18.6 kg/m<sup>2</sup> to 24.9 kg/m<sup>2</sup> | 0.18 | Ref | Ref |  |
| 25.0 kg/m<sup>2</sup> to 29.9 kg/m<sup>2</sup> | 0.28 | 1.61 (0.78 to 3.34) | +0.11 (-0.06 to +0.27) | 0.20 |
| More than 30.0 kg/m<sup>2</sup> | 0.24 | 1.38 (0.67 to 2.85) | +0.07 (-0.08 to +0.21) | 0.39 |
| **Hospitalized During Acute COVID Infection** |  |  |  |  |
| No | 0.19 | Ref | Ref |  |
| Yes | 0.38 | 1.97 (1.11 to 3.48) | +0.18 (+0.00 to +0.37) | 0.029 |
| **Symptom Count During Acute COVID Infection<sup>†,‡</sup>** |  |  |  |  |
| 0-5 symptoms | 0.21 | Ref | Ref |  |
| 6-8 symptoms | 0.21 | 1.00 (0.38 to 2.64) | +0.00 (-0.20 to +0.20) | 0.99 |
| 9-11 symptoms | 0.07 | 0.33 (0.07 to 1.53) | -0.14 (-0.31 to +0.04) | 0.15 |
| ≥ 12 symptoms | 0.25 | 1.21 (0.49 to 2.96) | +0.04 (-0.16 to +0.24) | 0.68 |
| **Self-reported Health Score at Worst Point in Acute COVID-19<sup>†,§</sup>** |  |  |  |  |
| >60 | 0.12 | Ref | Ref |  |
| 50-60 | 0.09 | 0.73 (0.11 to 4.69) | -0.03 (-0.24 to +0.17) | 0.74 |
| 30-49 | 0.21 | 1.68 (0.35 to 8.11) | +0.09 (-0.16 to +0.33) | 0.51 |
| <30 | 0.35 | 2.82 (0.66 to 12.1) | +0.23 (-0.05 to +0.51) | 0.14 |

\*Individuals whose BMI was <18.5 kg/m<sup>2</sup> (N=2) and who are Pacific Islander/Native Hawaiian (N=2) were omitted from the analyses
<sup>†</sup>Analyses only conducted amongst participants who were not hospitalized during their acute COVID-19 infection
<sup>‡</sup>N=132
<sup>§</sup>N=74

## DISCUSSION

Persistent symptoms among some persons who develop COVID-19 has led to the hypothesis that SARS-CoV-2 may, in some form or location, persist in the human host for long periods following acute infection.^19^ This hypothesis has motivated the search for SARS-CoV-2 far removed in time from acute infection and in sites disparate from those of acute infection.^8^ Overcoming many of the limitations of the initial work in this regard, we found evidence of SARS-CoV-2 in plasma for up to 14 months following SARS-CoV-2 infection and that this persistence is influenced by the events of acute infection.

Because our measurement of SARS-CoV-2 is via immunoreactivity, it cannot be assumed that every signal above background is specific for antigen from natural SARS-CoV-2 infection. Antigens from related pathogens or the host can theoretically cross-react. This differs from detection of nucleic acid for which specificity is more certain and more easily directly assessed by sequencing. Therefore, understanding the specificity of our Simoa® assay is critical for interpreting findings in SARS-CoV-2-infected persons, and this requires a true-negative group large enough to precisely estimate low-frequency false positivity. Using a diverse group of 250 adults sampled prior to the introduction of SARS-CoV-2 in humans, we found 2% false positivity in the assay. Of note, the quantitative values for the false positive specimens were well above limits of detection but not overtly related to any available demographic characteristic. This 98% specificity is high but not complete, meaning that caution would be needed if interpreting individual-level results in patients. Although not formally false positivity, there is also concern that vaccination against SARS-CoV-2 or recent reinfections could cloud interpretation of positive assay results.^8^ We mitigated this by studying specimens from an era that was largely prior to either of these occurrences and by detailed characterization of our participants.

Our inferences are consistent with most but not all prior smaller studies investigating SARS-CoV-2 persistence three or more months following acute infection. In plasma, Swank et al. (n=40 SARS-CoV-2-infected participants versus n=45 uninfected),^13^ Schultheiß et al. (n=29 versus n=2),^15^ and Craddock et al. (n=47 versus n=15)^16^ each found excess SARS-CoV-2 antigen prevalence in the post-acute period compared to uninfected persons, but Kanberg et al. (n=31 versus n=17) did not.^14^ Studies of tissue have featured even smaller numbers of participants,^8^ the exception being an investigation of 46 participants undergoing colonoscopy for inflammatory bowel disease in whom 70% had detectable SARS-CoV-2 RNA or antigen.^12^ The proportion of antigen positivity in our study was lower than most of the prior reports,^15,16^ including one using our assay.^13^ This may be explained by other studies being enriched with highly symptomatic patients recruited from Long COVID clinics whereas our population was consecutive volunteers who were interested in a variety of long-term aspects related to COVID-19. Alternatively, our work and the cited studies have used three different never-compared antigen assays that may not be interchangeable. In addition to direct detection of SARS-CoV-2 components in the post-acute phase of infection, evolving B cell immunity over time also indirectly suggests that some aspect of SARS-CoV-2 is persistent.^9^ Finally, regarding biologic plausibility, there is ample evidence of persistence of feline coronaviruses in their natural host.^20,21^

That persons hospitalized during acute COVID-19 had a significantly higher probability of persistent SARS-CoV-2 antigenemia, an observation also described by others,^16^ suggests the influence of the acute phase of infection in establishing the persistent SARS-CoV-2 reservoir. This was bolstered by the finding, although not meeting conventional statistical significance, that those reporting the most severe overall health during acute infection among the non-hospitalized also had substantially higher frequency of post-acute antigenemia. Coupled with the recent documentation of replication-competent virus in blood during acute infection,^22^ our findings suggest that SARS-CoV-2 might seed distal sites through the bloodstream and establish protected reservoirs in some sites. Alternatively, clinically more severe acute infection may simply be a marker of higher inoculum in the known sites of primary infection, which then have a greater chance of evading subsequent immune clearance. Our findings, however, provide no direct evidence regarding the persistent presence of replication-competent or even transcriptionally active virus. Yet, finding of circulating antigen so far in time from acute infection at least suggests some source of antigen production to counteract clearance. Whatever the mechanism for establishing a reservoir, our findings suggest an explanation for the protective effect of SARS-CoV-2 vaccination and antiviral therapy during acute infection against the occurrence of post-acute sequelae of COVID-19 (PASC).^23–25^

As in a prior report using our assay,^13^ antigen detection was intermittent over time in individuals in whom it was ever present. To what extent this represents true biologic variability versus lack of assay reproducibility is unknown. Formally, if true biologic variability, this could stem from variability in antigen production, release from reservoirs into the bloodstream, or clearance. A more sensitive assay that could interrogate larger plasma volumes, such as one being developed by our team,^26^ might provide substantial insights on these questions.

Our work has several limitations. First, our pandemic era group was a convenience sample, enriched to some unknown degree by those experiencing symptoms. Thus, the approximately 5% to 10% excess prevalence of antigenemia that we observed cannot be taken as a population-based estimate for all individuals experiencing SARS-CoV-2 infection during the period of our study. However, we suspect identifying a more representative group, replete with available blood specimens, will be highly challenging. Second, our participants were mainly infected with ancestral SARS-CoV-2 strains without the benefit of prior vaccination or effective anti-viral agents. It is thus unclear how our findings will generalize to contemporary infections. Finally, while our participants did not have known or clinically suspected reinfections prior to antigen detection, they were not systematically assessed for asymptomatic SARS-CoV-2 infection. It is therefore possible that some fraction of the antigenemia is from recent re-infection instead of the distant primary infection.

In summary, our data provide among the strongest evidence to date that SARS-CoV-2 should be added to the list of RNA viruses whose components may persist beyond the period of acute illness. Given the scale of the pandemic and knowledge that SARS-CoV-2 spike is highly immunogenic,^27–29^ there are now urgent questions regarding whether persistent SARS-CoV-2 is causally related to either the chronic symptoms of PASC (e.g., fatigue, pain, and cognitive difficulty) or discrete incident complications (e.g., cardiovascular events). Addressing this agenda will require observational as well as concurrent experimental studies that target inhibition of viral component production, neutralization or clearance of existing antigen, and/or modulation of the immune response to these antigens. Such studies are now underway (NCT05877507, NCT05595369, NCT05576662, NCT05668091, and NCT06161688).

## Data Availability

All data produced in the present study are available upon reasonable request to the authors.

## FOOTNOTES

## Acknowledgements

We are grateful to the study participants and their medical providers. We acknowledge current LIINC clinical study team members Grace Anderson, Kofi Asare, Melissa Buitrago, Emily Conway, Marin Ewing, Emily Fehrman, Tony Figueroa, Diana Flores, Marian Kerbleski, Antonio Rodriguez, Justin Romero, Dylan Ryder, Matthew So, Viva Tai, Alex Tang, and Meghann Williams; and current LIINC laboratory team members Amanda Buck, Brian LaFranchi, and David Maison. We thank Jessica Chen, Aidan Donovan, Carrie Forman, and Rania Ibrahim for assistance with data entry and review. We thank the UCSF AIDS Specimen Bank for processing specimens and maintaining the LIINC biospecimen repository. We are grateful to Elnaz Eilkhani and Monika Deswal for regulatory support. We also acknowledge all former LIINC team members.

## Author Contributions

Designed the study: MJP, SGD, DRW, JNM

Designed the cohort: MJP, PYH, MSD, TJH, JDK, SGD, JNM

Collected biospecimens and clinical data: MJP, RH, ML, CCS, AC, AZ, ML, BH, KA.

Performed the assays: ZS, EB, YS in the laboratory of DRW.

Managed the data: SL, TD, BV.

Analyzed the data: MJP, SAG, DVG.

Wrote the manuscript: MJP, SGD, JDK, DRW, JNM.

Edited the manuscript: All authors.

Approved the manuscript: All authors.

---

**Supplemental Table 1.** Differences in prevalence of SARS-CoV-2 antigen positivity in plasma among participants in the post-acute phase of COVID-19 in comparison to pre-pandemic participants. P-values represent chi-square and Fisher’s 2-sided exact test as appropriate.

**Any SARS-CoV-2 Antigen**
| **Group** | **No. of participants** | **No. (%) antigen positive** | **Prevalence difference 95% confidence interval; p value** |
| --- | --- | --- | --- |
| Pre-pandemic | 250 | 5 (2.0%) | Reference |
| Pandemic |  |  |  |
| 3 to 6.0 months | 151 | 19 (13%) | +0.11 (+0.050 to +0.16); p < 0.001 |
| 6 to 10.0 months | 131 | 14 (11%) | +0.087 (+0.031 to +0.14); p < 0.001 |
| 10 to 14 months | 122 | 9 (7.4%) | +0.054 (+0.0042 to +0.10); p = 0.017 |

**Spike**
| **Group** | **No. of participants** | **No. (%) antigen positive** | **Prevalence difference 95% confidence interval; p value** |
| --- | --- | --- | --- |
| Pre-pandemic | 250 | 3 (1.2%) | Reference |
| Pandemic |  |  |  |
| 3 to 6.0 months | 151 | 9 (6.0%) | +0.048 (+0.0075 to +0.088; p = 0.012) |
| 6 to 10.0 months | 131 | 7 (5.3%) | +0.041 (+0.0006 to +0.082; p = 0.036) |
| 10 to 14 months | 122 | 5 (4.1%) | +0.029 (-0.0087 to +0.067; p = 0.12) |

**Nucleocapsid**
| **Group** | **No. of participants** | **No. (%) antigen positive** | **Prevalence difference 95% confidence interval; p value** |
| --- | --- | --- | --- |
| Pre-pandemic | 250 | 2 (0.80%) | Reference |
| Pandemic |  |  |  |
| 3 to 6.0 months | 151 | 7 (4.6%) | +0.038 (+0.0031 to +0.074; p = 0.030) |
| 6 to 10.0 months | 131 | 3 (2.3%) | +0.015 (-0.013 to +0.043; p = 0.34) |
| 10 to 14 months | 122 | 2 (1.6%) | +0.0084 (-0.017 to +0.034; p = 0.60) |

**S1**
| **Group** | **No. of participants** | **No. (%) antigen positive** | **Prevalence difference 95% confidence interval; p value** |
| --- | --- | --- | --- |
| Pre-pandemic | 250 | 3 (1.2%) | Reference |
| Pandemic |  |  |  |
| 3 to 6.0 months | 151 | 4 (2.7%) | +0.015 (-0.015 to +0.043; p = 0.43) |
| 6 to 10.0 months | 131 | 4 (3.1%) | +0.019 (-0.014 to +0.051; p = 0.24) |
| 10 to 14 months | 122 | 3 (2.5%) | +0.013 (-0.018 to +0.043; p = 0.40) |

